# Prevalence of Hospital PCR Confirmed COVID-19 Cases in Patients with Chronic Inflammatory and Autoimmune Rheumatic Diseases

**DOI:** 10.1101/2020.05.11.20097808

**Authors:** José L. Pablos, Lydia Abasolo-Alcázar, José M. Álvaro-Gracia, Francisco J. Blanco, Ricardo Blanco, Isabel Castrejón, David Fernández-Fernández, Benjamín Fernández-Gutierrez, María Galindo, Miguel A. González-Gay, Sara Manrique-Arija, Natalia Mena-Vázquez, Antonio Mera-Varela, Miriam Retuerto, Álvaro Seijas-Lopez, RIER investigators group

## Abstract

**Background:** The susceptibility of patients with rheumatic diseases, and the risks or benefits of immunosuppressive therapies for COVID-19 are unknown.

**Methods:** We performed a retrospective study with patients under follow-up in rheumatology departments from seven hospitals in Spain. We matched updated databases of rheumatology patients with SARS-CoV-2 positive PCR tests performed in the hospital to the same reference populations. Rates of PCR+ confirmed COVID-19 were compared among groups.

**Results:** Patients with chronic inflammatory diseases had 1.32-fold higher prevalence of hospital PCR+ COVID-19 than the reference population (0.76% vs 0.58%). Systemic autoimmune or immune mediated diseases (AI/IMID) patients showed a significant increase, whereas inflammatory arthritis (IA) or systemic lupus erythematosus (SLE) patients did not. COVID-19 cases in some but not all diagnostic groups had older ages than cases in the reference population. IA patients on targeted-synthetic or biological disease-modifying antirheumatic drugs (ts/bDMARD), but not those on conventional-synthetic (csDMARD), had a greater prevalence despite a similar age distribution.

**Conclusion:** Patients with AI/IMID show a variable risk of hospital diagnosed COVID-19. Interplay of aging, therapies, and disease specific factors seem to contribute. These data provide a basis to improve preventive recommendations to rheumatic patients and to analyze the specific factors involved in COVID-19 susceptibility.

## INTRODUCTION

The severe lung and systemic inflammatory manifestations observed in severe acute respiratory distress syndrome-coronavirus-2 (SARS-CoV-2) infection, have led to the hypothesis of a hyperinflammatory mechanism, more dependent on the host response than on direct viral cellular damage (1,2). Certain parallelisms with other cytokine storm situations, such as macrophage activation syndrome (MAS) or chimeric antigen receptor T cells (CAR-T) associated systemic inflammatory syndromes have been invoked in support. This hypothesis has prompted the rapid introduction of anti-inflammatory and immunomodulatory agents approved for rheumatic diseases in the therapeutic strategies to combat SARS-CoV-2 infection.

The prevalence of severe SARS-CoV-2 infection in patients with previous autoimmune or inflammatory diseases is unknown. This group of patients is not represented in the largest Chinese series as a specific risk factor for susceptibility or severity of COVID-19 (3,4). An excess of morbimortality associated to the previous use of conventional or targeted immunosuppressive drugs has neither been reported. However, in an Italian series of 1591 severe, ICU admitted COVID-19 patients, the most prevalent comorbidity in patients under 40 years-old was a miscellanea of patients that included inflammatory and immunosuppressed patients (5).

A potential preventive or therapeutic effect of certain immunomodulatory therapies in these patients has been hypothesized. Among them, antimalarials, colchicine, corticosteroids, jakinibs and IL-6 or IL1 antagonists are being used under special conditions or clinical trials with weak evidences (6). However, the risks of these drugs in the context of viral infections without concomitant antiviral therapies are not negligible (7-9). Whether immunosuppressants put patients with rheumatic disease at an increased or decreased risk for severe COVID-19 is unknown, and evidence is urgently needed to guide prevention and therapy (10,11).

Since timely obtaining methodologically rigorous data on the prevalence of severe SARS-CoV-2 infection in our patients under different therapies is challenging at this moment (8), we have performed an exploratory analysis of the relative prevalence of hospital diagnosed COVID-19 in large multicentric cohorts of rheumatic patients under follow-up.

## PATIENTS AND METHODS

We performed a retrospective observational study with patients under follow-up in rheumatology departments from reference hospitals in Spain. We selected seven centers pertaining to a public research network for the investigation of inflammation and rheumatic diseases (RIER), by the availability of updated medical records ID lists of adult patients under follow-up in rheumatology departments, diagnosed of chronic inflammatory arthritis (IA) or systemic autoimmune or immune-mediated conditions (AI/IMID) including: rheumatoid arthritis (RA), psoriatic arthritis (PSA), spondyloarthritis (SpA), systemic lupus erythematosus (SLE), Sjögren’s syndrome (SS), systemic sclerosis (SSc), systemic lupus erythematosus (SLE), polymyalgia rheumatica or giant cell arteritis (PMR-GCA) and other diseases (including systemic vasculitis, Behcet’s syndrome, sarcoidosis, and inflammatory myopathies). We also obtained updated ID lists of patients with IA on therapy with only conventional synthetic disease-modifying antirheumatic drugs (csDMARD), specifically methotrexate or leflunomide, or with targeted synthetic or biologic ts/bDMARD, including jakinibs or any biological agent. Not all groups have databases of all diagnostic or therapeutic categories. Only complete datasets allowing to calculate incident/total number of cases in a particular category were included for the aggregated analysis.

Selected centers were also microbiology reference centers where all SARS-CoV-2 PCR diagnostic tests in the covered adult population were performed. Patients’ medical record ID’s were matched against central SARS-CoV-2+ PCR hospital registers between April 7^th^ and 17^th^, after the incidence peak of SARS-CoV-2 infection had been reached in Spain. Medical records were reviewed to confirm clinical COVID-19 diagnosis. Since SARS-CoV-2 PCR availability was limited, these registries only include patients attending referral hospitals, and exclude the less severe community cases that did not require hospitalization nor referral to hospitals emergency departments.

Data are reported as rates in the different groups and compared with the rates in the general reference population by odds ratios (OR) with 95% confidence intervals (CI) and chi-square tests. Median age in the different groups was compared by non-parametric tests.

## RESULTS

The reference population in the participating hospitals was 2.9 million people, with a global prevalence of PCR+ positive SARS-CoV-2 of 0.58%. In the different centers, it varied from 0.23% to 1.16%, consistently with the reported prevalence in the different regions of Spain (https://cnecovid.isciii.es/covid19/). We screened a total of 26.131 patients under follow-up in rheumatology departments for hospital positive SARS-CoV-2 PCR results in their reference hospitals, and found a higher prevalence of PCR+ cases (0.76%; OR 1.3, CI 1.15-1.52) compared to the reference population, with a similar regional variation from 0.23% to 1.24%. Although we cannot calculate the age-adjusted rates due to lack of age information of PCR-negative cases, PCR+ cases in a representative sample (n 3,800) of the reference population had a median younger age than rheumatic disease cases (55 vs 65 years old, p<0.0001). Age and sex distribution are shown in Table 1.

**Table 1.**
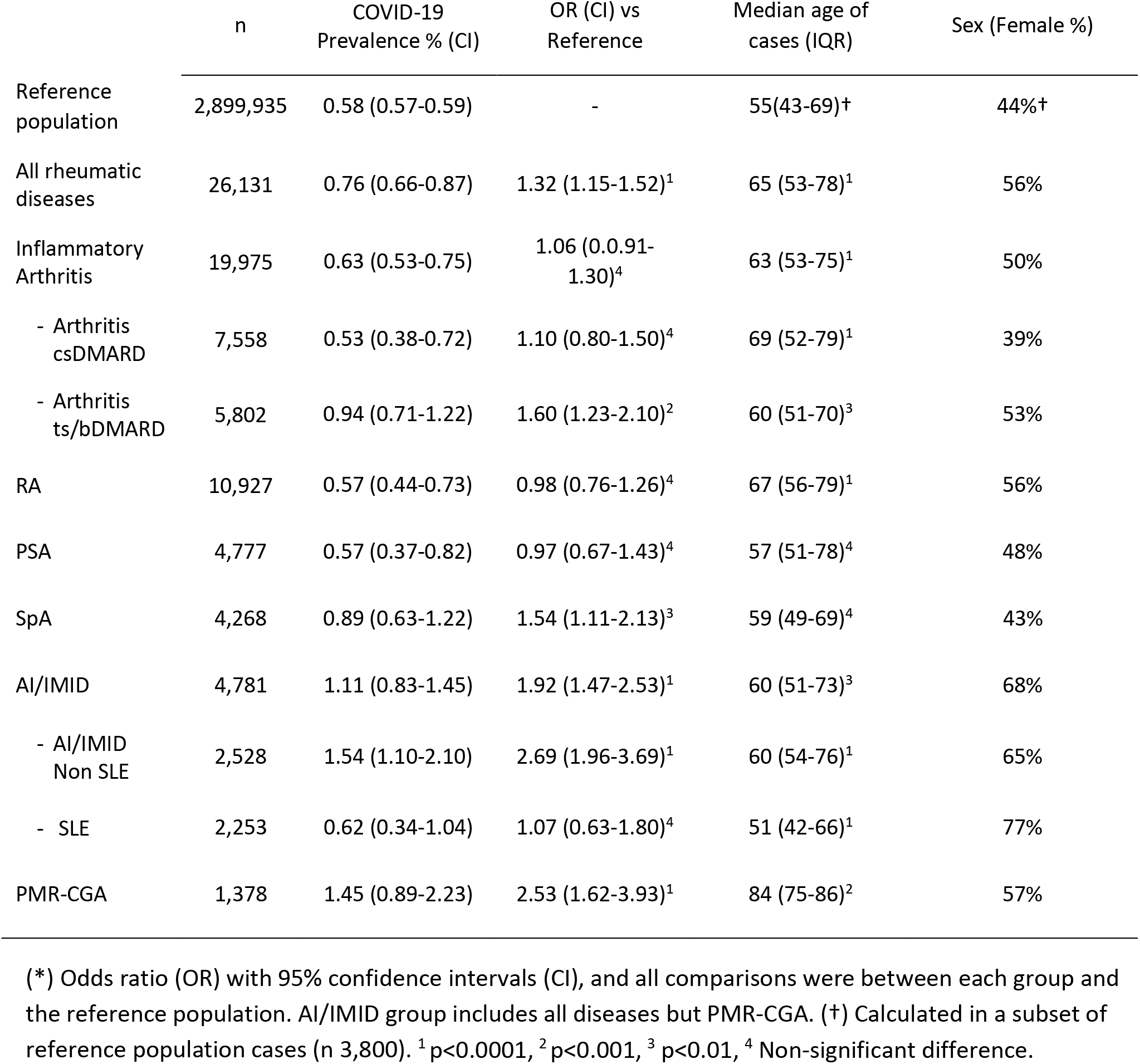
Rates of hospital PCR+ confirmed COVID-19 cases in patients with different rheumatic diseases.

Patients with IA (RA, PSA and SpA) showed a prevalence similar to that in the reference population, but in the SpA subset in was increased by 1.54-fold (CI 1.11-2.13). Among this group, patients with IA on therapy with csDMARD (methotrexate or leflunomide) also showed a similar prevalence, whereas patients on ts/bDMARD therapy showed a 1.60-fold (CI 1.23-2.10) increased prevalence of COVID-19 compared to the reference population (Table 1 and Figure 1).

**FIGURE 1.**
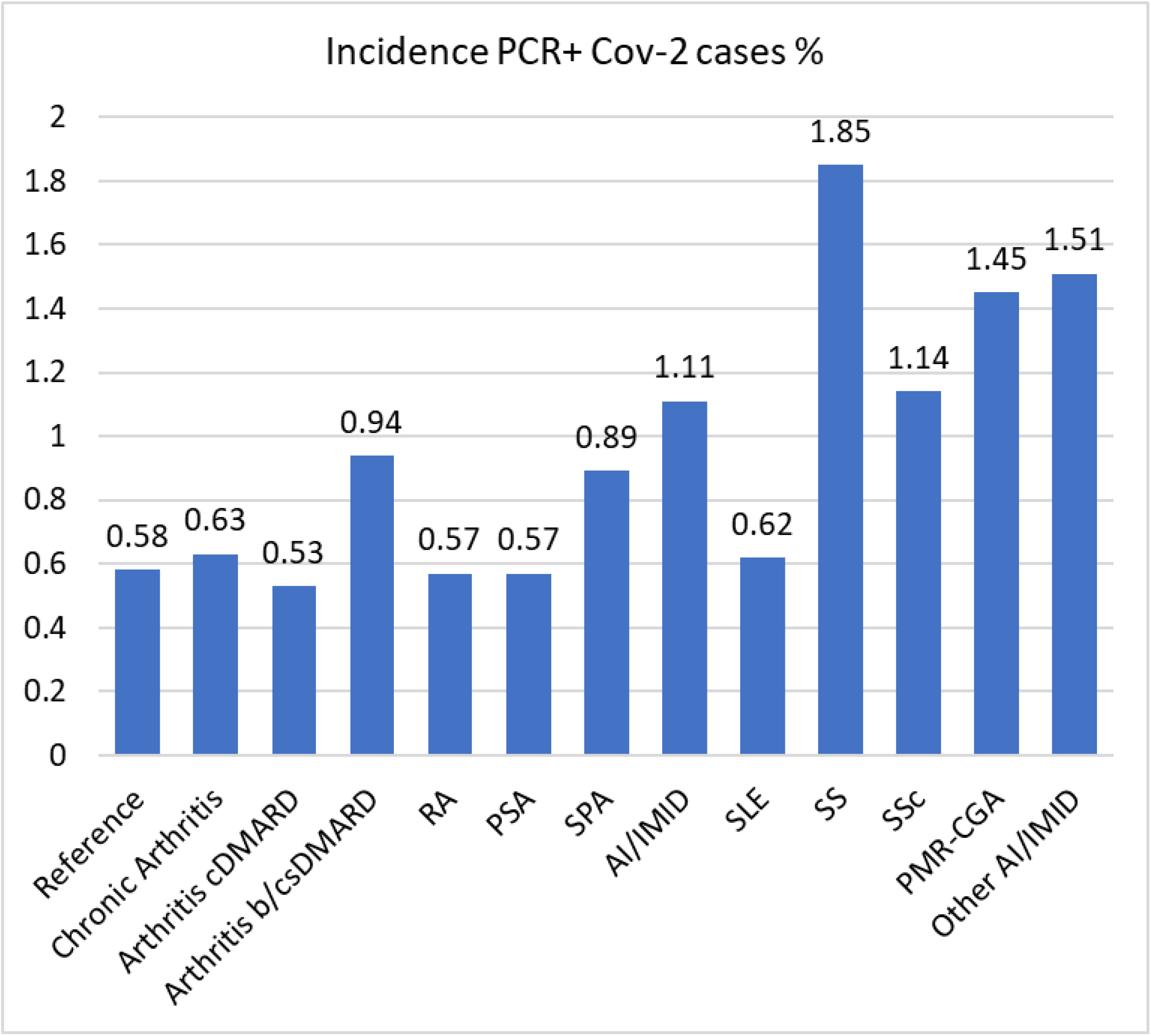
Rates of hospital COVID-19 in patients with chronic arthritis, autoimmune diseases and reference population. Prevalence of hospital PCR confirmed cases of COVID-19 infection in patients with chronic IA or AI/IMID diseases (n 26,131) in seven reference hospitals in Spain, compared to that in the reference population of the same hospitals (n 2.9 million). AI/IMID group include all diagnosis but PMR-CGA, and Other AI/IMID a of the less frequent diseases as indicated in Patients and Methods.

All aggregated groups of patients with AI/IMID showed higher rates of COVID-19, and analyses of the different groups confirmed increased prevalence in all diagnostic groups but SLE, where it was remarkably lower than in the other groups and similar to the reference population (Table 1 and Figure 1).

Regarding the age distribution of COVID-19 cases in the different IA and AI/IMID, some but not all diagnostic groups were older compared to the reference population (Table 1). The age of PCR-negative cases was not available in all cohorts. A partial sample from two centers showed that the proportion of rheumatic patients older than 65 doubled that in the reference population (40% vs 20%). The relative prevalence in older (>65 years) versus younger individuals was similar in rheumatic patients (OR 1.77; CI 1.20-2.60; n 8,270) and in the reference population (OR 1.86; CI 1.73-1.99; n 372,000).

## DISCUSSION

Our systematic approach identified a significant number of patients with different rheumatic conditions and immunomodulatory therapies with SARS-CoV-2 PCR confirmed diagnosis that allow us to describe the prevalence of hospital COVID-19 and to identify differences between diagnostic and therapeutic groups. The observed differences can be considered to identify specific factors associated to susceptibility to COVID-19 in rheumatic patients.

Patients with RA or PSA showed similar rates compared to the reference population despite their older age, but specific groups of IA including patients with SpA and those on ts/bDMARD seem at higher risk. This suggests that these specific immunomodulators may increase the risk for COVID-19, similarly as for other viral infections (7,12).

In patients with different systemic AI/IMID, a variably greater prevalence among specific disease groups was detected. It is remarkable the relatively low rate in SLE patients, despite an expected greater use of corticosteroids and immunosuppressants. A possible explanation is the frequent use of antimalarials which might have played a protective role as proposed according to their *in vitro* antiviral effects, but this will require further analyses (13). This relatively low prevalence contrasts with the significant increase observed in all other AI/IMID diseases. In most groups, aging is an expected associated risk-factor but somewhat surprisingly, in the PMR-GCA group, hospital COVID-19 prevalence does not seem higher compared to other groups of AI/IMID. The real risk in PMR-CGA might have been underestimated, considering the difficult access to the hospital of aged and institutionalized patients and merits further analysis.

Several considerations have to be taken to interpret these results. We have only identified cases requiring attention in hospital emergency departments and often hospitalization. Since severity, but not prevalence, increases with age, a higher rate of severe cases among rheumatic patients’ cohorts was expected (14,15). However, only in some groups, hospital COVID-19 cases have an older age than the reference population. Therefore, although older age is a clear risk factor, disease or therapy related factors also seem to modify the risk for hospital COVID-19 cases in the different rheumatic patients’ groups. PCR testing bias in rheumatic patients is unlikely because during the peak phase, authorities’ recommendations for hospital attendance were based on severity rather than on pre-existing potential risk conditions. We are aware of a still unknown but important number of rheumatic patients with milder SARS-CoV-2 disease whose detection was based on self-reporting and phone consultations, but not confirmed by PCR testing. The real prevalence of severe and non-severe cases among rheumatic patients, but also in the non-rheumatic reference population, could be only reliably estimated by future serologic testing.

With these limitations, we conclude that the different chronic inflammatory or autoimmune diseases patients and therapies have a different impact on the risk COVID-19. Our data on specific groups should be translated to current recommendations regarding alert on infection risk and prevention measures in rheumatic patients. Ongoing studies of the specific factors potentially involved in the observed differences will hopefully contribute to understand the impact of the SARS-CoV-2 pandemics in different risk groups.

## Data Availability

Most data are included in the manuscript. Additional data can be shared upon reasoned request to the corresponding author.

## ACKNOWLEDGEMENTS

We are grateful to Rafael Delgado (Hospital 12 de Octubre), Luis Salas (Hospital General Universitario Gregorio Marañón), Begoña Palop-Borrás (Hospital Regional Universitario de Málaga) and all Microbiology and Infectious Diseases Departments of participating centers for providing registers of PCR results, and to Celia Iglesias for valuable help in the preparation of the manuscript.

## COMPETING INTERESTS

None of the authors have competing interests to declare.

## ETHICAL APPROVAL INFORMATION

The study was approved by Comité de Ética de la Investigación del Hospital Universitario 12 de Octubre, CEIm number: 20/160.

## FUNDING INFORMATION

The RIER network was supported by the Fondo de Investigación Sanitaria, Instituto de Salud Carlos III (RD16/0012 RETICS program) and cofinanced by the European Regional Development Fund (FEDER).

## CONTRIBUTORSHIP

JLP and MG take responsibility for the integrity of the data, data analysis and statistical analyses. All authors participated in acquisition of data, designing the analyses, interpreting the results and writing the manuscript. RIER investigators participated in the design and partially collaborated in acquisition of data.

## DATA SHARING STATEMENT

All data relevant to the study are included in the article. Data are available upon reasonable request to corresponding author.

## PATIENT AND PUBLIC INVOLVEMENT

Patients and/or the public were not involved in the design, or conduct, or reporting or dissemination plans of this research.

## KEY MESSAGES

What is already known about this subject?

- The susceptibility of patients with rheumatic diseases, and the risks or benefits of immunosuppressive therapies for COVID-19 are unknown.

What does this study add?

- This study shows that there is an increased rate of hospital COVID-19 cases associated to some but not all chronic inflammatory or autoimmune conditions compared to the reference population.
- The risks seem to depend on age, the specific disease, and previous therapies, remarkably increased rates were observed in biologic or targeted synthetic but not in conventional DMARD treated patients.

How might this impact on clinical practice or future developments?

- These data may be translated to current recommendations regarding alert on infection risk and prevention measures in rheumatic patients.
- The observed differences will hopefully contribute to future studies to identify the risk factors for COVID-19 in different patients’ groups.

## Notes

### Competing Interest Statement

The authors have declared no competing interest.

### Funding Statement

The study was performed by the RIER research network which was supported by the Fondo de Investigacion Sanitaria, Instituto de Salud Carlos III (RD16/0012 RETICS program), and cofinanced by the European Regional Development Fund (FEDER).

### Author Declarations

The study was approved by Comite de Etica de la Investigacion del Hospital Universitario 12 de Octubre, CEIm number: 20/160.

